# Three Distinct Trajectories of Red Blood Cell Distribution Width and Their Significant Association with Mortality in Sepsis Patients: A Group-Based Trajectory Modeling Study with Validation

**DOI:** 10.64898/2026.02.25.26347114

**Authors:** Lei Cai, Yuwei Hua, Wei Lu, Haozhen Bing, Qinfen Gao, Wei Zhang

## Abstract

The red cell distribution width (RDW) is a recognized prognostic marker in sepsis, yet its dynamic changes over time and their relationship with outcomes remain unexplored. This study aimed to identify distinct RDW trajectories during the early phase of sepsis and evaluate their association with mortality. We conducted a retrospective cohort study using data from the MIMIC-IV database (n=3,813) as the derivation cohort and from the First Affiliated Hospital of Kunming Medical University (n=467) for external validation. Sepsis patients with at least seven RDW measurements within the first ten days of hospitalization were included. Group-based trajectory modeling (GBTM) was employed to identify RDW trajectories. A three-trajectory model was selected based on model fit indices and clinical interpretability: Trajectory 1 (Slow-Decrease, 32.97%), Trajectory 2 (Slow-Increase, 43.30%), and Trajectory 3 (Fluctuating-Rapid Decrease, 23.73%). In the our study, Cox models adjusted for confounders revealed that, compared to Trajectory 1, Trajectory 3 was independently associated with significantly increased 30-day (HR 1.47, 95% CI 1.17–1.84) and 90-day mortality (HR 1.54, 95% CI 1.25–1.88). Conversely, Trajectory 2 was associated with the most favorable survival rates. Kaplan-Meier analysis consistently showed the highest mortality in the Trajectory 3 group. External validation confirmed the model’s robustness and the consistent prognostic value of the identified trajectories. We conclude that dynamic RDW trajectories, readily identifiable from routine clinical data, provide significant prognostic information beyond single-time-point measurements and can aid in the risk stratification of sepsis patients.

## 1 Introduction

Sepsis is a life-threatening organ dysfunction caused by a dysregulated host response to infection. It poses a major threat to human life and safety. Among the 48.9 million sepsis cases reported globally in 2017, the mortality rate was approximately 22.5%, accounting for nearly 20% of global deaths that year[1]. Concurrently, sepsis is one of the leading causes of in-hospital mortality, with a fatality rate of 18.7% among hospitalized sepsis patients, a figure that can reach up to 55.7% in the Intensive Care Unit (ICU) [2]. Despite established frameworks like Sepsis-3 and the Surviving Sepsis Campaign, accurately predicting outcomes for sepsis patients remains challenging.

The Red Blood Cell Distribution Width (RDW) is a common parameter in a complete blood count, indicating the variability in the size of circulating red blood cells. Historically, RDW was primarily used for the diagnosis and differential diagnosis of iron-deficiency anemia and other chronic anemias. Recent studies, however, have found that elevated RDW is associated with increased mortality in various conditions, including septic cardiomyopathy[3], acute myocardial infarctionp[4-5], heart failure[6-7], cardiac arrest[8], and acute pulmonary embolism[9]. Nevertheless, previous research has predominantly analyzed RDW at single, fixed time points, overlooking the relationship between its dynamic changes and patient prognosis in sepsis. Furthermore, as a readily available component of the routine complete blood count used clinically, RDW facilitates easy serial monitoring. This study aims to identify distinct RDW trajectories in sepsis patients using Group-Based Trajectory Modeling (GBTM), a method that accommodates unbalanced panels and missing data.

## 2 Methods

### 2.1 Data sources

Patient data for the modeling cohort were extracted from the MIMIC-IV database, version 3.1 (application number: 66772901). The validation cohort consists of inpatients admitted to the First Affiliated Hospital of Kunming Medical University (KMUFAH) from 2018 to 2025. The MIMIC-IV database is a publicly available electronic health database containing clinical data from approximately 300,000 patients admitted to the Beth Israel Deaconess Medical Center (BIDMC) in Boston between 2008 and 2019. The database has been approved by the Institutional Review Board (IRB) of the Massachusetts Institute of Technology (MIT). All protected health information has been de-identified; specific records (e.g., dates of birth, admission, and discharge) were encrypted to protect patient privacy, thus granting an exemption from further ethical review. The data from the First Affiliated Hospital of Kunming Medical University has been approved by the Hospital Ethics Committee. This retrospective study was conducted in accordance with the Declaration of Helsinki. Due to the retrospective nature of the study and the use of anonymized data (for MIMIC-IV) and data collected from routine clinical care with waived consent approved by the ethical committee (for KMUFAH data), the requirement for informed consent was waived by the respective IRBs/Ethics Committees (MIT IRB and the Ethics Committee of the First Affiliated Hospital of Kunming Medical University).”

### 2.2 Study Population

The inclusion criteria were as follows: (i) meeting the Sepsis-3.0 diagnostic criteria (confirmed or suspected infection plus an increase in Sequential Organ Failure Assessment [SOFA] score ≥ 2 points); (ii) age ≥ 18 years; and (iii) having at least seven RDW measurements within the first ten days of hospitalization. The exclusion criteria were: (i) a history of comorbidities including malignant tumors, hematological diseases, chronic obstructive pulmonary disease (COPD), obstructive sleep apnea, acute or chronic blood loss, or blood transfusion; and (ii) excessive missing data. The detailed patient screening process is shown in Figure 1.

**Figure 1.**
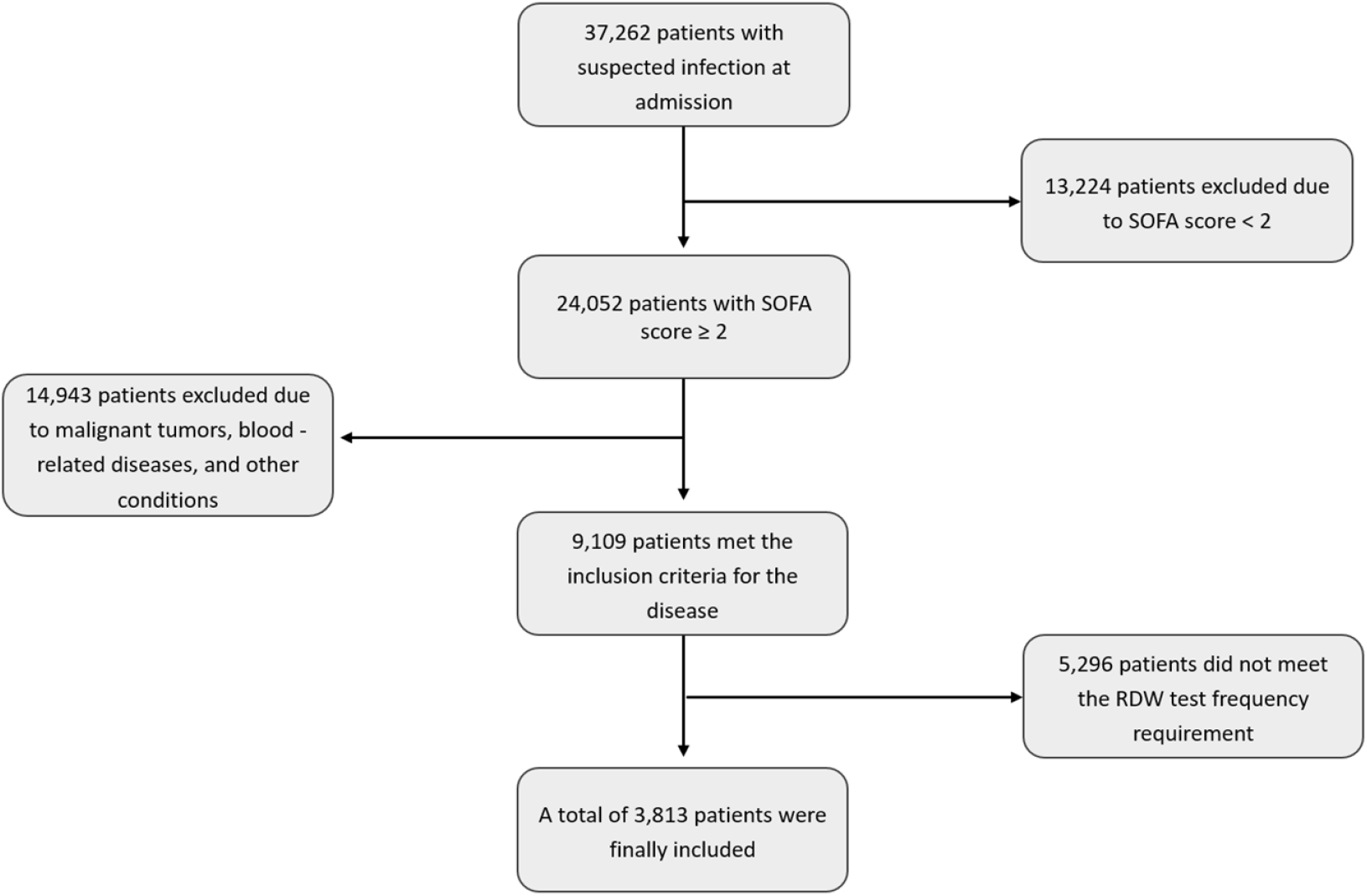
Flowchart of Patient Selection from the MIMIC-IV Database

### 2.3 Clinical Data

The study extracted the following information from the MIMIC-IV database for eligible patients for statistical analysis: basic demographic data, diagnoses, outcomes, survival time, RDW values and their corresponding measurement times, ICU length of stay, Sequential Organ Failure Assessment (SOFA) score, Glasgow Coma Scale (GCS) score, Acute Physiology Score III (APS III), Oxford Acute Severity of Illness Score (OASIS), Model for End-Stage Liver Disease (MELD) score, and Charlson Comorbidity Index (CCI).

### 2.4 Statistical Analysis

Group-based trajectory modeling (GBTM) was employed to identify distinct RDW trajectories. The optimal number of trajectory groups was selected based on the smallest Akaike Information Criterion (AIC) and Bayesian Information Criterion (BIC), combined with clinical interpretability. Given the minimal amount of missing data, median imputation was used. Each patient was assigned to a trajectory group based on the model’s posterior probability. The Kolmogorov-Smirnov test was used to assess the normality of data distribution. Normally distributed continuous data are presented as mean ± standard deviation and were compared using one-way analysis of variance (ANOVA). Non-normally distributed continuous data are presented as median (interquartile range) and were compared using the Kruskal-Wallis H test. The Kaplan-Meier method was used to plot survival curves, and Cox proportional hazards models were applied to analyze the impact of different covariates on survival. All statistical analyses were performed using R software (version 4.4.2).

## 3 Results

### 3.1 Group-Based Trajectory Modeling

We employed Group-Based Trajectory Modeling (GBTM) to identify distinct RDW trajectories. Based on the minimization criteria for Akaike Information Criterion (AIC) and Bayesian Information Criterion (BIC), along with clinical interpretability, a 3-trajectory model was selected (Figure 2). Trajectory 1 (Traj 1): Slow-Decrease group, included 1257 patients (32.97%), characterized by a gradual decrease in RDW. Trajectory 2 (Traj 2): Slow-Increase group, included 1651 patients (43.30%), characterized by a gradual increase in RDW. Trajectory 3 (Traj 3): Fluctuating-Rapid Decrease group, included 905 patients (23.73%), characterized by a baseline RDW value above normal levels, which fluctuated initially followed by a rapid decrease.

**Figure 2.**
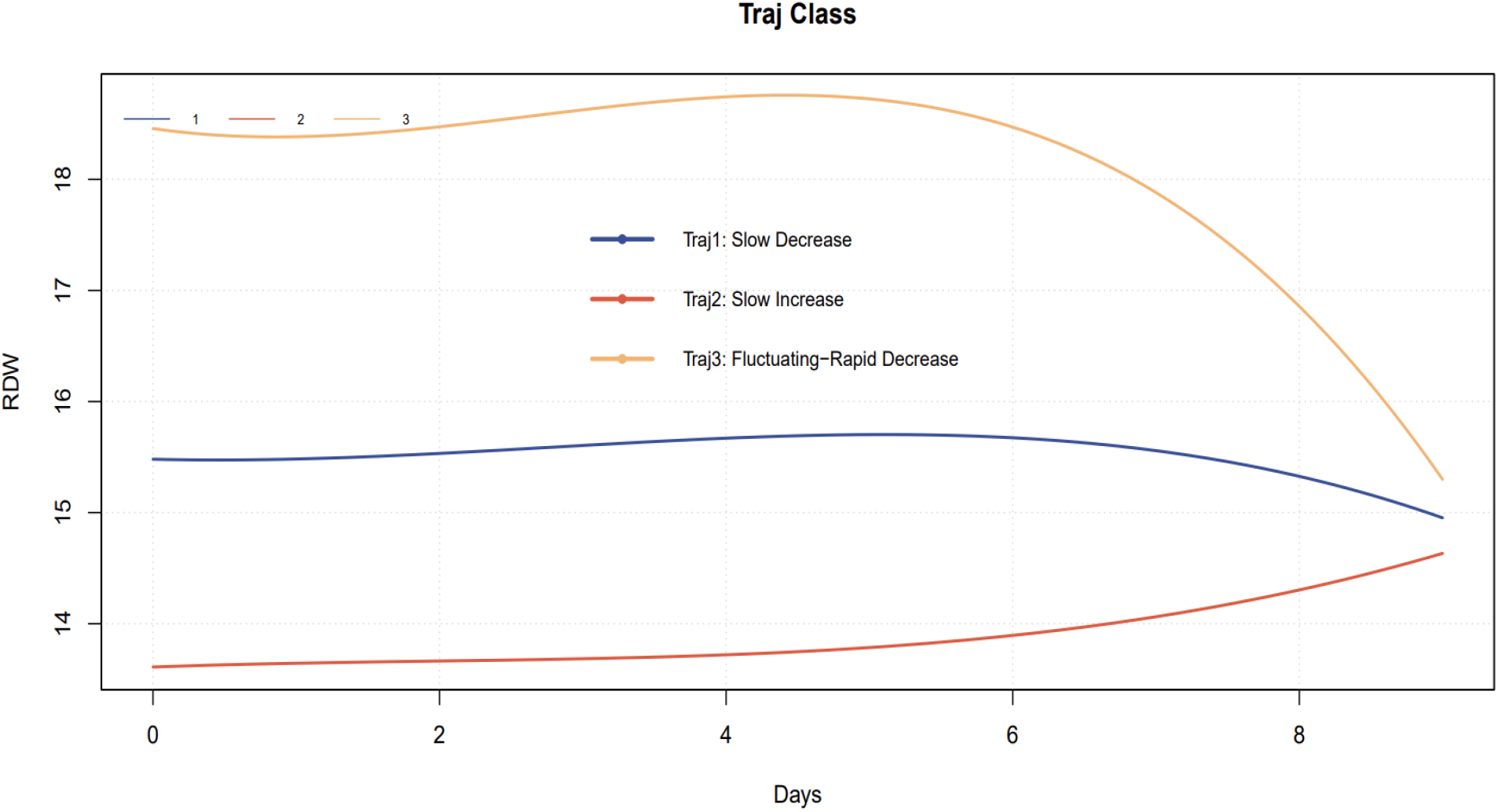
Identified RDW Trajectories in Sepsis Patients

**Figure 3.**
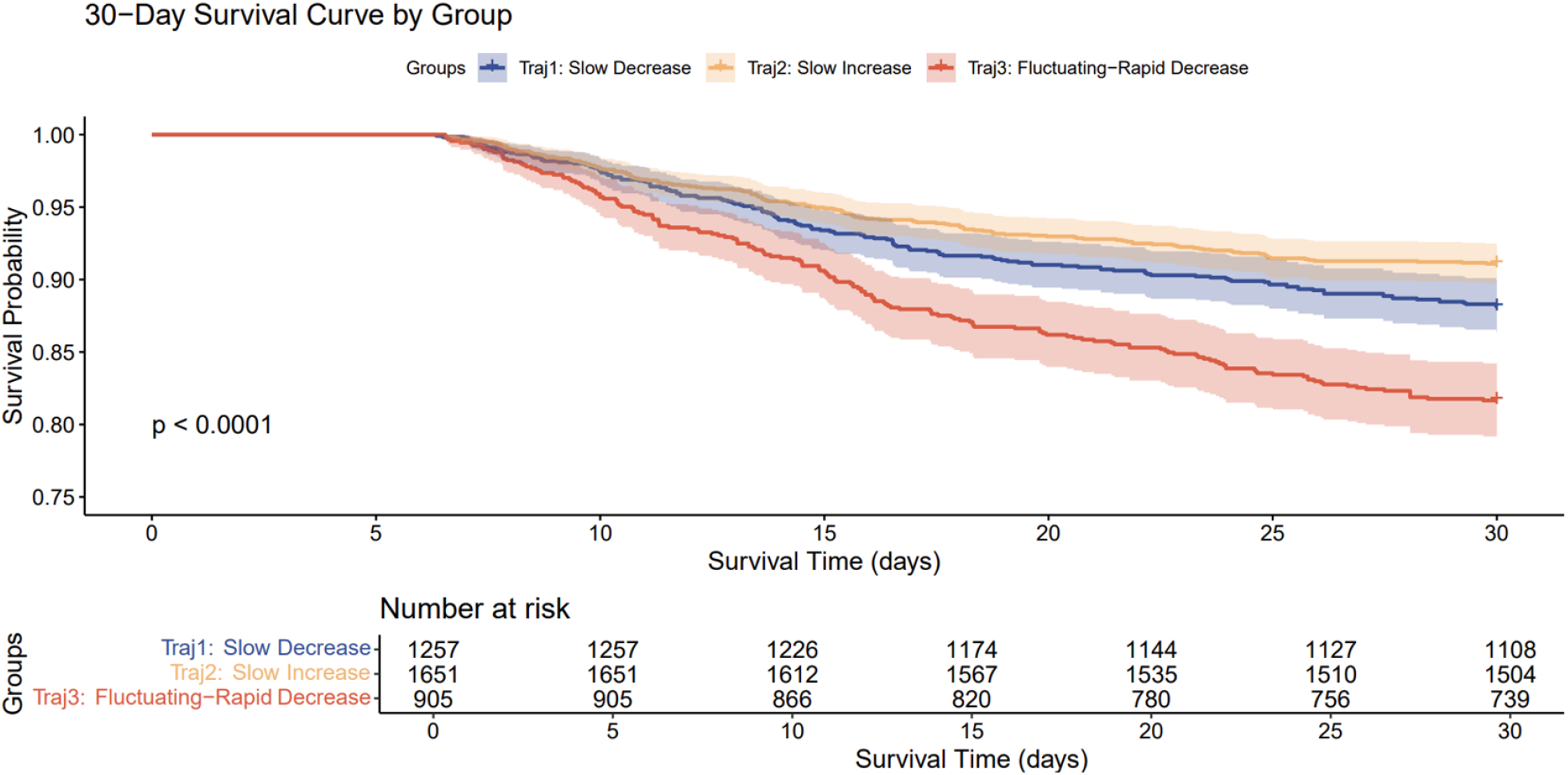
30-day Kaplan-Meier Survival Curves for Patients with Different Trajectories

**Figure 4.**
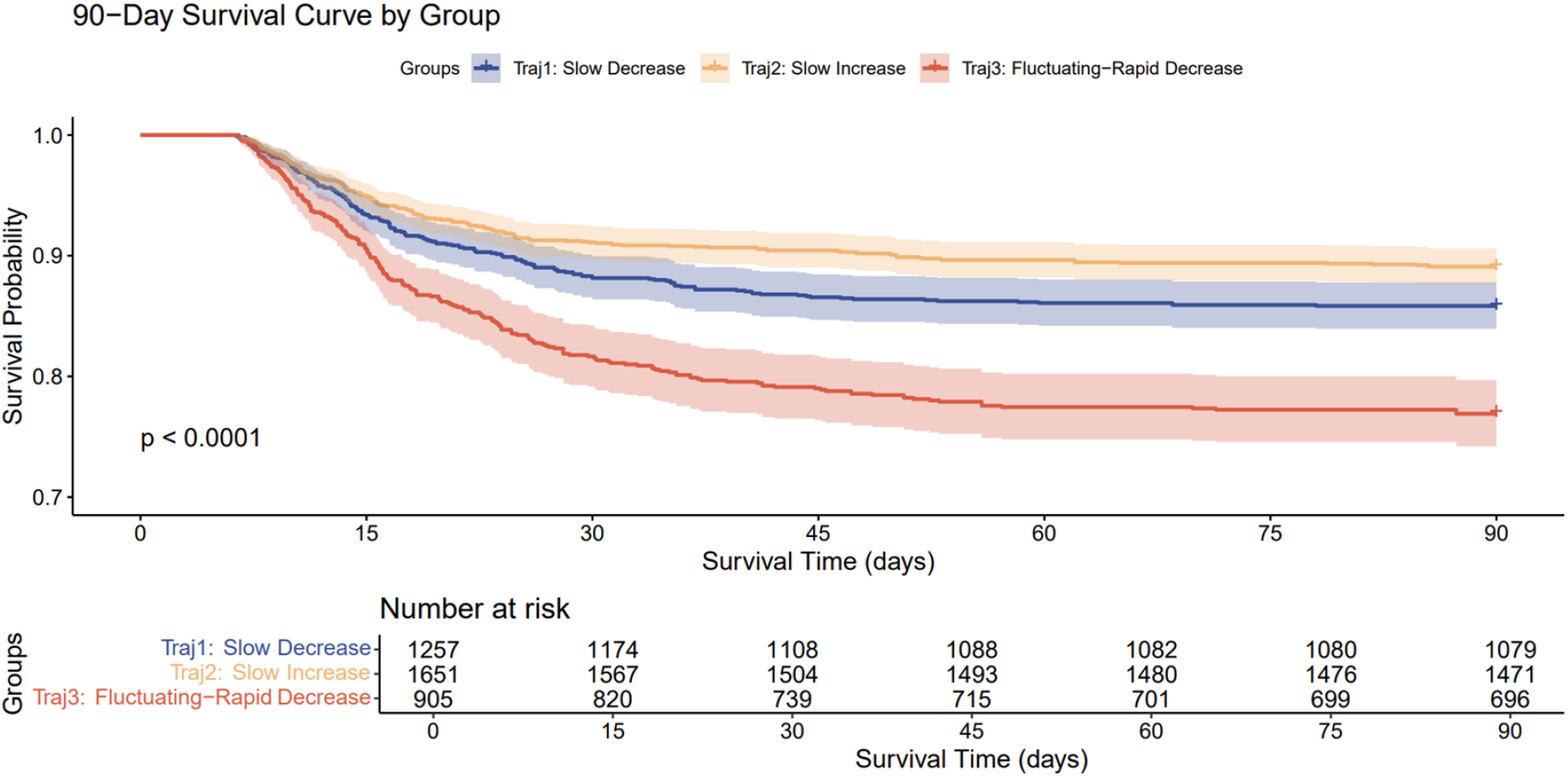
90-day Kaplan-Meier Survival Curves for Patients with Different Trajectories

### 3.2 Baseline Characteristics of the Patients

Patients were assigned to the Traj 1 (n=1257), Traj 2 (n=1651), and Traj 3 (n=905) groups based on the maximum posterior probability. The Kruskal-Wallis H test indicated significant differences in baseline characteristics among the different trajectory groups. Patients in the Traj 3 group had the highest SOFA, APS III, and MELD scores, alongside the highest in-hospital, 30-day, and 90-day mortality rates. Regarding ICU stay, the Traj 3 group had the shortest length of stay. Conversely, the Traj 2 group had the longest hospital and ICU lengths of stay, and the lowest APS III scores, in-hospital mortality, 30-day mortality, and 90-day mortality rates

**Table 1.** Clinical Characteristics of Patients with Different RDW Trajectories.

|  | Traj1 (n=1257) | Traj 2 (n=1651) | Traj 3 (n=905) | P |
| --- | --- | --- | --- | --- |
| Age (years) | 66.0 (54.0, 77.0) | 66.0 (53.0, 77.0) | 65.0 (55.0, 76.0) | *P* = 0.466 |
| Sex(male) | 641 (51.0%) | 886 (53.7%) | 448 (49.5%) | *P* = 0.103 |
| Hospital LOS (days) | 14.5 (9.9, 23.3) | 15.4 (10.1, 25.4) | 14.9 (10.4, 25.9) | *P* = 0.028 |
| ICU LOS (days) | 4.7 (2.3, 10.0) | 5.8 (2.8, 11.9) | 4.0 (2.0, 9.2) | *P* < 0.001 |
| SOFA | 3.0 (2.0, 4.0) | 3.0 (2.0, 4.0) | 4.0 (2.0, 5.0) | *P* < 0.001 |
| GCS | 15.0 (14.0, 15.0) | 15.0 (14.0, 15.0) | 15.0 (14.0, 15.0) | *P* = 0.009 |
| APSIII | 52.0 (41.0, 66.0) | 46.0 (34.0, 58.0) | 57.0 (46.0, 71.0) | *P* < 0.001 |
| OASIS | 35.0 (29.0, 41.0) | 34.0 (28.0, 39.0) | 35.0 (29.0, 41.0) | *P* < 0.001 |
| MELD | 15.0 (10.0, 21.0) | 11.0 (8.0, 16.0) | 20.0 (12.0, 25.0) | *P* < 0.001 |
| SIRS | 3.0 (2.0, 3.0) | 3.0 (2.0, 3.0) | 3.0 (2.0, 3.0) | *P* = 0.110 |
| Charlson Comorbidity Index | 6.0 (4.0, 8.0) | 5.0 (3.0, 7.0) | 6.0 (5.0, 8.0) | *P* < 0.001 |
| In-hospital mortality | 183 (14.6%) | 181 (11.0%) | 217 (24.0%) | *P* = 0.121 |
| 30-day mortality | 149 (11.9%) | 147 (8.9%) | 166 (18.3%) | *P* < 0.001 |
| 90-day mortality | 178 (14.2%) | 180 (10.9%) | 209 (23.1%) | *P* < 0.001 |

Data are presented as median (interquartile range) or n (%). **LOS**, length of stay; **SOFA**, Sequential Organ Failure Assessment; **GCS**, Glasgow Coma Scale; **APS III**, Acute Physiology Score III; **OASIS**, Oxford Acute Severity of Illness Score; **MELD**, Model for End-Stage Liver Disease; **SIRS**, Systemic Inflammatory Response Syndrome.

### 3.3 Cox Proportional Hazards Models and Kaplan-Meier Survival Curves

Cox proportional hazards models and Kaplan-Meier survival curves were used to analyze the association between different RDW trajectories and mortality. In the unadjusted model, compared to the Traj 1 group, the Traj 3 group was significantly associated with increased 30-day (HR 1.60, 95% CI 1.29-2.00, P<0.001) and 90-day mortality (HR 1.71, 95% CI 1.40-2.09, P<0.001). Conversely, the Traj 2 group was significantly associated with decreased 30-day (HR 0.74, 95% CI 0.59-0.93, P=0.010) and 90-day mortality (HR 0.76, 95% CI 0.62-0.93, P=0.008). Even after adjusting for confounding factors, the Traj 3 group remained significantly associated with increased 30-day and 90-day mortality, while the Traj 2 group remained significantly associated with decreased 30-day and 90-day mortality. Consistent with the Cox model results, the Kaplan-Meier survival curves demonstrated that the Traj 3 group had the highest mortality rates at both 30 and 90 days, whereas the Traj 2 group had the lowest mortality rates at both time points.

**Table 2.**
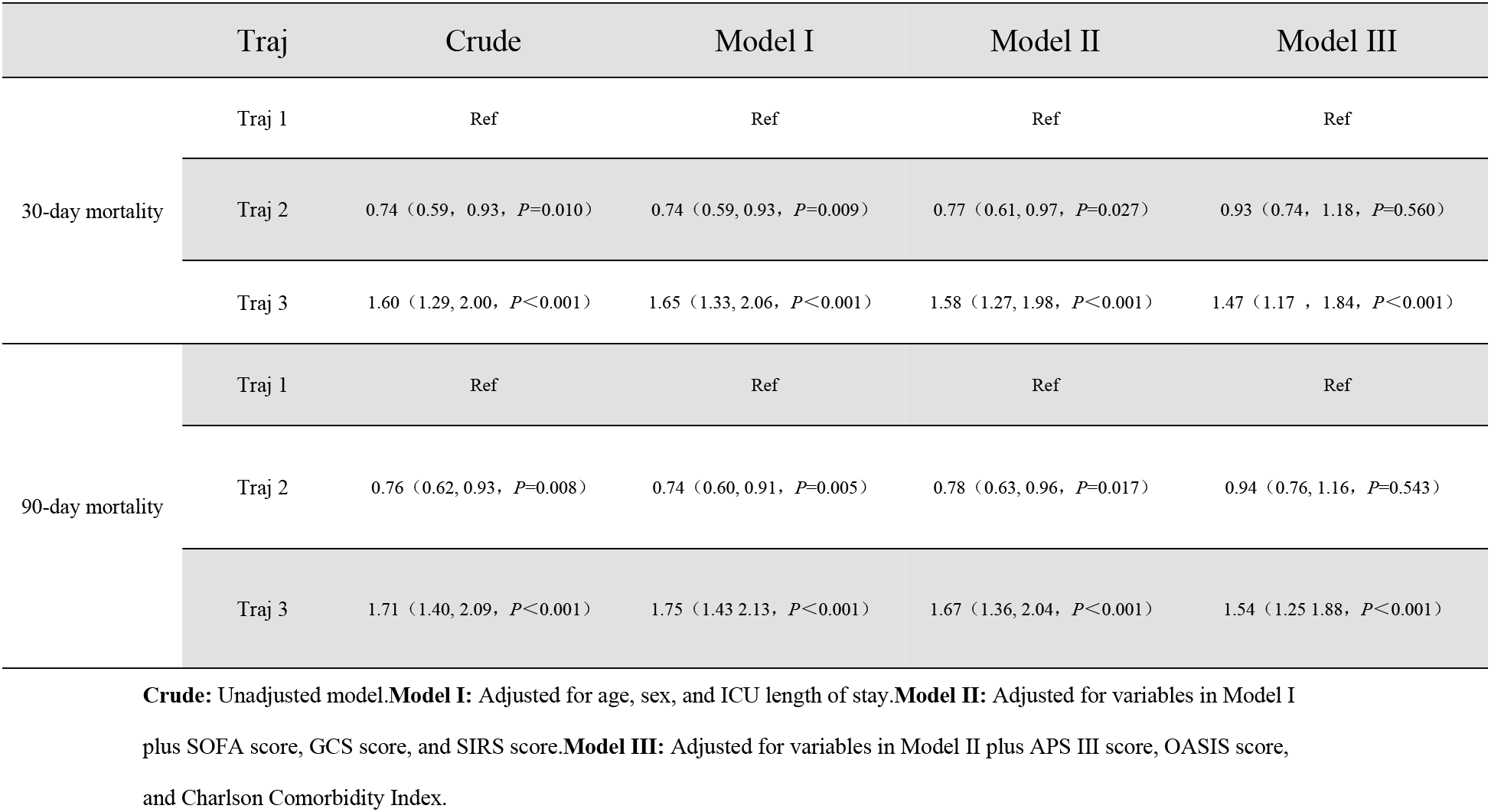
Univariable and Multivariable Cox Proportional Hazards Models for Different RDW Trajectories.

### 3.4 External Validation

We additionally collected clinical data from sepsis patients hospitalized at the First Affiliated Hospital of Kunming Medical University between 2018 and 2025. This dataset included detailed information such as basic demographics, diagnoses, outcomes, treatment regimens, and laboratory results, encompassing a total of 11,336 patients. After applying the same inclusion and exclusion criteria used for the modeling cohort, 467 patients were included in the validation study. We similarly employed GBTM to identify RDW trajectories in these patients. The resulting trajectory model closely resembled that derived from the MIMIC-IV database (as shown in Figure 5), also comprising three distinct trajectory groups: Trajectory 1 (Slow-Decrease group), Trajectory 2 (Slow-Increase group), and Trajectory 3 (Fluctuating-Rapid Decrease group).

**Figure 5.**
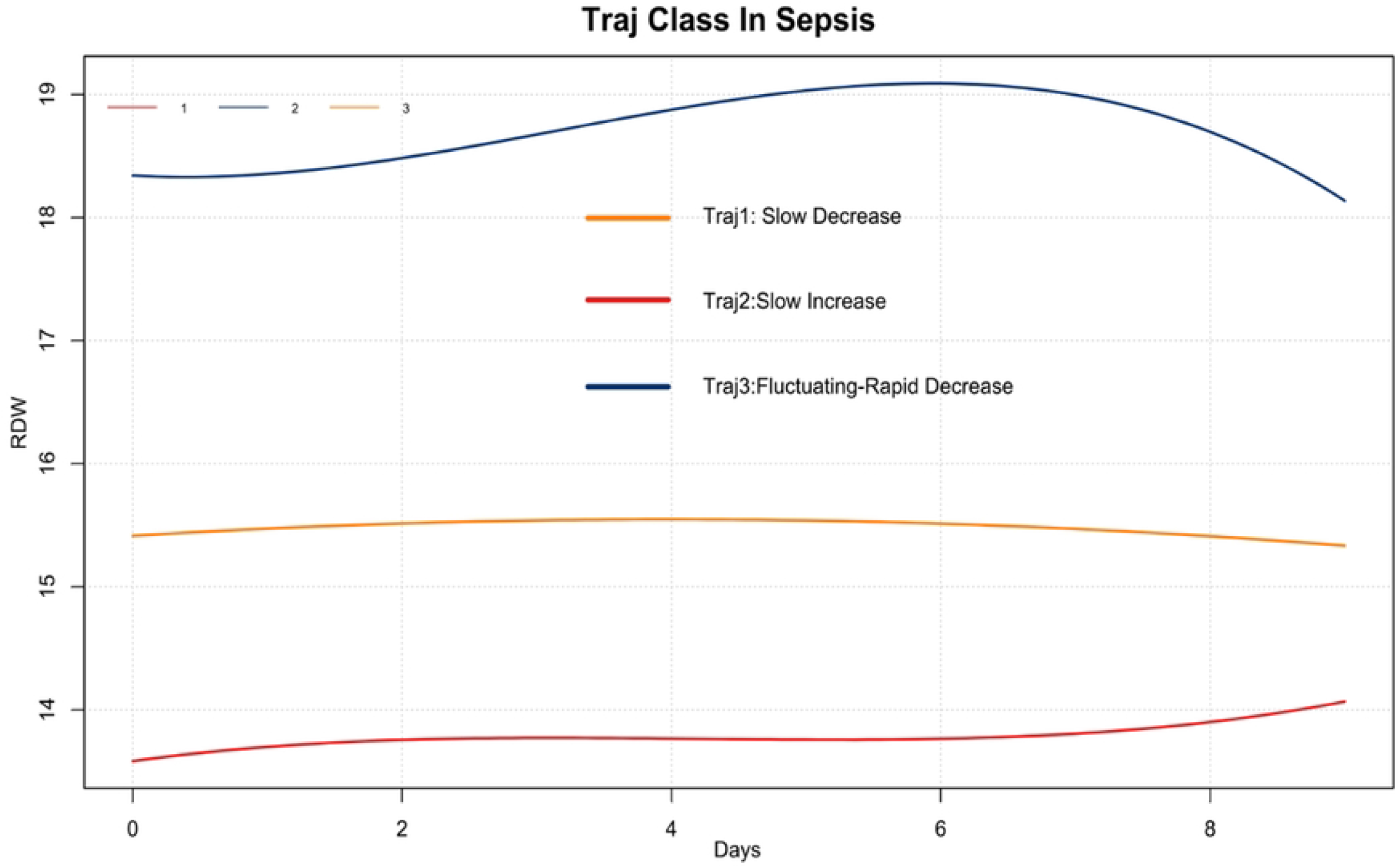
Validation of Trajectories Using Data from the First Affiliated Hospital of Kunming Medical University

**Figure 6.**
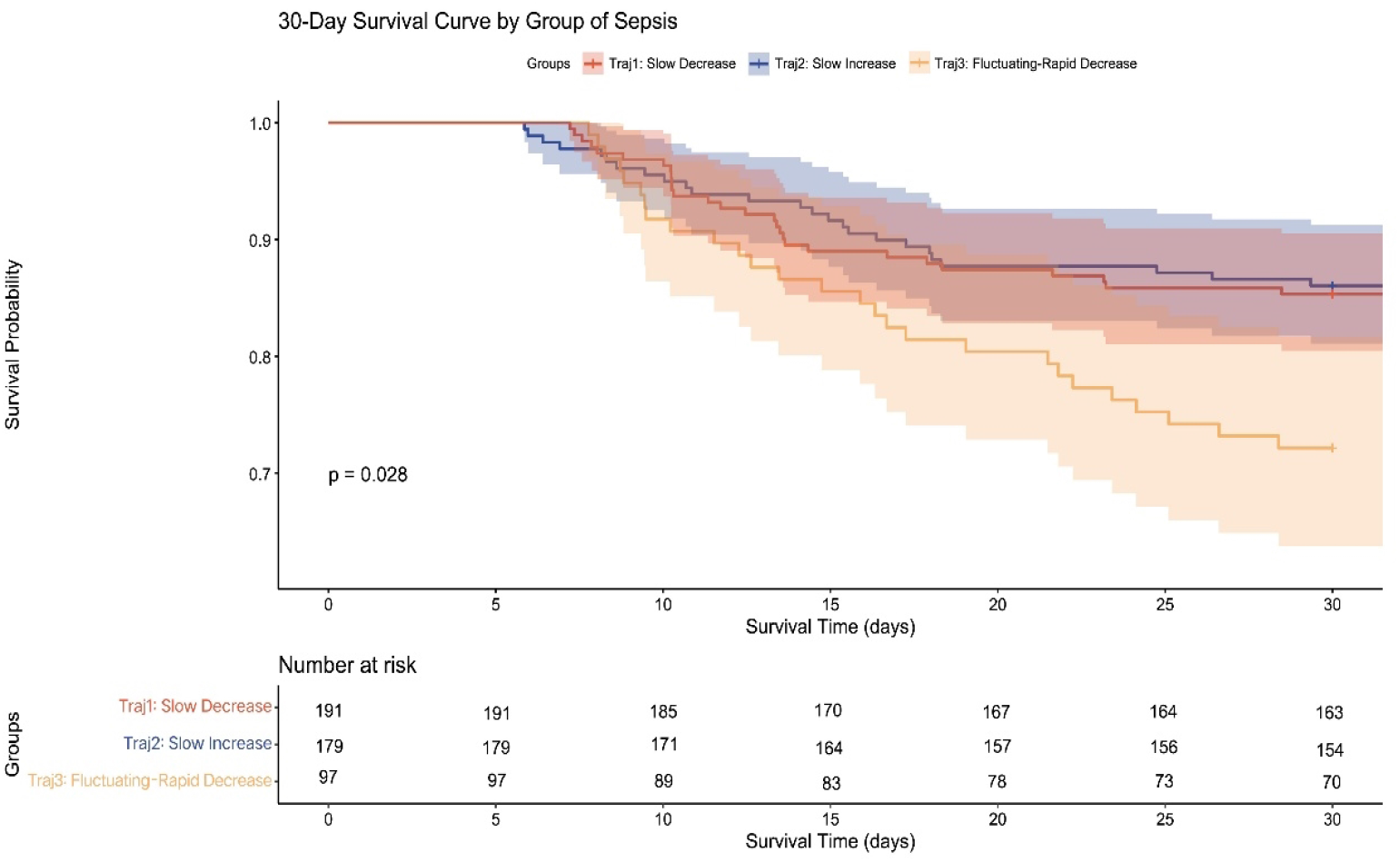
Kaplan-Meier Survival Curves for Different Trajectories Derived from the First Affiliated Hospital of Kunming Medical University Data

After assigning patients to groups based on the maximum posterior probability, we plotted Kaplan-Meier survival curves for 30-day survival for the three groups. The results indicated that mortality in the Traj 3 group was substantially higher than in the other two groups, which was consistent with our model’s predictions. Pairwise Log-rank tests between the three different trajectories revealed that both the survival rate and survival time of Traj 3 patients were significantly different from those of Traj 1 and Traj 2 patients (*P*=0.017 and *P*=0.019, respectively), while no significant difference was observed between Traj 1 and Traj 2 (*P*=0.793).

## Discussion

Our study yielded several notable findings. In this research, we initially employed Group-Based Trajectory Modeling (GBTM) to identify distinct RDW trajectories in sepsis patients. Based on the Akaike Information Criterion (AIC), Bayesian Information Criterion (BIC), and clinical interpretability, we selected a 3-trajectory model. Patients were subsequently assigned to one of the three distinct trajectory groups based on the maximum posterior probability. Using Trajectory 1 (Slow-Decrease group) as the reference, we performed Cox proportional hazards regression and Kaplan-Meier survival analysis. Furthermore, we validated the trajectory model using a local dataset from our hospital, which confirmed the model’s robustness. In our established model, the Traj 3 group had the highest SOFA scores and initial RDW values and was associated with the worst 30-day and 90-day outcomes. Conversely, the Traj 2 group had the longest ICU length of stay and the lowest initial RDW values, and was associated with the most favorable 30-day and 90-day prognosis. To the best of our knowledge, this is the first study to dynamically analyze the trajectory of RDW changes throughout the course of sepsis.

RDW is an indicator reflecting the variability in size of circulating red blood cells. Erythropoietin (EPO) is the hormone with the most significant influence on RDW. Abnormal production of EPO and reduced responsiveness of erythroid precursors to EPO can both lead to an elevated RDW[10], with the latter playing a more substantial role in sepsis. Inflammatory cytokines such as TNF-α, IL-1β, and IL-6 can decrease the sensitivity of bone marrow erythroid cells to EPO, leading to the release of immature red blood cells into the circulation, thereby increasing ineffective erythropoiesis and raising RDW. Studies have shown that the negative impact of sepsis on the hematopoietic system is manifested by the fact that the hyperinflammatory state during sepsis reduces red blood cell lifespan[10], increases erythrocyte heterogeneity, impairs normal red blood cell production, and consequently leads to an increase in RDW[3,10-13]. On the other hand, the inflammatory state promotes the expression of hepcidin via the JAK-STAT3 pathway[14]. Hepcidin is a central regulator of iron metabolism; it binds to the iron exporter ferroportin, inducing its degradation, thereby inhibiting iron efflux and leading to impaired iron utilization. During inflammation, hepcidin blocks iron release from macrophages and iron absorption in the gut, resulting in hypoferremia and anemia of inflammation, causing ineffective erythropoiesis and heterogeneous red cell sizes, which elevates RDW[15]. Conversely, sepsis is often accompanied by hypoxia. Hypoxia can directly suppress hepcidin production through the HIF/Transmembrane Serine Protease 6 (TMPRSS6)/Membrane-type Serine Protease-2 (MT2) pathway[16], promoting iron release and accelerating erythropoiesis, albeit potentially with reduced uniformity. In summary, the alteration of RDW in sepsis results from the interplay of multiple factors, including inflammation, impaired iron utilization, and hypoxia.

RDW not only holds diagnostic value in chronic inflammatory conditions but recent studies have also demonstrated its role in the diagnosis and prognosis prediction of acute inflammatory diseases like sepsis. For instance, Kim et al. found that sepsis patients whose RDW increased within 72 hours of emergency department admission had a higher mortality risk[17]. Wang et al. discovered that RDW level is an independent predictor of mortality in elderly sepsis patients in the ICU, with each 1% increase in RDW associated with an 18% increase in mortality[15]. Gu et al. reported that an elevated RDW-to-albumin ratio (RAR) was significantly associated with increased in-hospital mortality in sepsis patients with atrial fibrillation[18]. Zeng et al. found that an increased RDW was associated with in-hospital, short-term, and long-term mortality in critically ill patients with atrial fibrillation[19]. Han YQ et al. demonstrated that elevated RDW was associated with poor long-term outcomes in sepsis patients, and RDW showed higher predictive accuracy compared to five traditional severity scoring systems (APS III, MLODS, OASIS, SOFA, SIRS, SAPS II, and qSOFA)[20]. In another study by Kim YC et al., a prognostic model for sepsis constructed using increased RDW (>14.5%), elevated delta neutrophil index (DNI) (>5.0%), or decreased platelet count (<150,000/mm^3^) demonstrated good predictive ability (AUC=0.758) [21]. Ku NS et al. found that the RDW level at the onset of bacteremia was an independent predictor of 28-day mortality[22]. Kim CH et al. showed that an increase in RDW from baseline within the first 72 hours of hospitalization could serve as a strong independent predictor of mortality in patients with severe sepsis or septic shock[23]. Notably, Fontana V et al., by monitoring sublingual microcirculation, found no relationship between changes in RDW and microcirculatory alterations in septic patients[24]. In our study, after adjusting for confounding factors including peripheral vascular disease in the Cox model, analysis showed that it was not a risk factor for patient mortality, which is consistent with the conclusions of Fontana V et al.

In a previous prospective multicenter study, researchers found that RDW values during the first week of ICU stay were consistently higher in non-surviving sepsis patients compared to survivors. Furthermore, significant associations were observed between RDW and levels of oxidative stress (MDA) and inflammation (TNF-α)[25]. However, these studies primarily focused on RDW values at specific time points, overlooking the patterns of dynamic change. In the present study, we identified different RDW trajectories using GBTM and validated the model’s rationality and scientific merit using a local dataset. Based on previous reports, we hypothesize that distinct RDW trajectories may reflect different immune phenotypes in sepsis patients. In the Traj 3 group, which had the highest SOFA scores, initial RDW values, and mortality, the initially elevated RDW may indicate a state of immune overactivation. The subsequent rapid decline after fluctuation could signify a transition into an immunosuppressed state. Conversely, the slowly increasing RDW in the Traj 2 group, which had the lowest initial RDW and mortality, might suggest the absence of sepsis-induced immune dysregulation. This could explain the highest mortality in the Traj 3 group and the lowest in the Traj 2 group, although this hypothesis warrants further investigation.

The management of sepsis patients primarily involves three aspects: source control, hemodynamic stabilization, and modulation of the host immune response. Specifically, for patients in the Traj 3 group, early administration of anti-inflammatory agents, such as corticosteroids, might be more beneficial. As the disease progresses, the observation of a rapid decrease in RDW could indicate a need for immunostimulatory therapies, such as thymosin α1, which might help improve outcomes[26-28].Our trajectory model may provide a novel tool for identifying specific patient subgroups that could benefit from immunomodulatory therapy, offering a more precise approach than blanket treatment for all sepsis patients.

This study has limitations. The requirement for multiple RDW measurements at different time points to construct the trajectories necessitated the inclusion of patients who had frequent RDW tests. These patients often had more severe conditions requiring close monitoring, potentially leading to the exclusion of patients with milder infections. However, RDW is one of the most readily available clinical parameters. It closely reflects the patient’s inflammatory status and is considerably less expensive than other inflammatory markers (e.g., cytokines). Dynamic monitoring of RDW trajectories and identifying the trajectory type could assist clinicians in early assessment of patient condition, adjusting treatment strategies, ultimately improving patient outcomes, and reducing the burden of sepsis-related hospitalization.

In conclusion, using GBTM, we identified three distinct RDW trajectories in sepsis patients. Traj 2 was associated with higher 30-day and 90-day survival rates, while Traj 3 was associated with higher 30-day and 90-day mortality. These findings may aid clinicians in patient stratification and prognosis assessment.

## Data Availability

Data cannot be shared publicly due to restrictions imposed by the ethical review board and institutional policies designed to protect patient confidentiality. The data underlying the results presented in this study are available from the Ethics Committee of the First Affiliated Hospital of Kunming Medical University for researchers who meet the criteria for access to confidential data. Researchers may submit data access requests along with a detailed research proposal to the

https://physionet.org/content/mimiciv/2.2/

## Data Availability

The datasets analyzed during the current study are available from the MIMIC-IV database, upon completion of the required training and application. The clinical data from the First Affiliated Hospital of Kunming Medical University that were used for validation are not publicly available due to patient privacy and ethical restrictions but are available from the corresponding author on reasonable request.

